# Spot the Difference: Can ChatGPT4-Vision Transform Radiology Artificial Intelligence?

**DOI:** 10.1101/2023.11.15.23298499

**Authors:** Brendan S Kelly, Sophie Duignan, Prateek Mathur, Henry Dillon, Edward H Lee, Kristen W Yeom, Pearse Keane, Aonghus Lawlor, Ronan P Killeen

## Abstract

OpenAI’s flagship Large Language Model ChatGPT can now accept image input (GPT4V). “Spot the Difference” and “Medical” have been suggested as emerging applications. The interpretation of medical images is a dynamic process not a static task. Diagnosis and treatment of Multiple Sclerosis is dependent on identification of radiologic change. We aimed to compare the zero-shot performance of GPT4V to a trained U-Net and Vision Transformer (ViT) for the identification of progression of MS on MRI.

170 patients were included. 100 unseen paired images were randomly used for testing. Both U-Net and ViT had 94% accuracy while GPT4V had 85%. GPT4V gave overly cautious non-answers in 6 cases. GPT4V had a precision, recall and F1 score of 0.896, 0.915, 0.905 compared to 1.0, 0.88 and 0.936 for U-Net and 0.94, 0.94, 0.94 for ViT.

The impressive performance compared to trained models and a no-code drag and drop interface suggest GPT4V has the potential to disrupt AI radiology research. However misclassified cases, hallucinations and overly cautious non-answers confirm that it is not ready for clinical use. GPT4V’s widespread availability and relatively high error rate highlight the need for caution and education for lay-users, especially those with limited access to expert healthcare.

**Key points:**

- Even without fine tuning and without the need for prior coding experience or additional hardware, GPT4V can perform a zero-shot radiologic change detection task with reasonable accuracy.
- We find GPT4V does not match the performance of established state of the art computer vision models. GPT4V’s performance metrics are more similar to the vision transformers than the convolutional neural networks, giving some possible insight into its underlying architecture.
- This is an exploratory experimental study and GPT4V is not intended for use as a medical device.

**Summary statement:** GPT4V can identify radiologic progression of Multiple Sclerosis in a simplified experimental setting. However GPT4V is not a medical device and its widespread availability and relatively high error rate highlight the need for caution and education for lay-users, especially those with limited access to expert healthcare.

## Introduction

Multiple sclerosis (MS) is a chronic inflammatory, demyelinating neurodegenerative disease of the Central Nervous System (CNS)(1). Magnetic resonance imaging (MRI) is the most important tool for diagnosis and surveillance due to its high sensitivity for the assessment of inflammatory and neurodegenerative changes in the CNS(2). New and enlarging lesions are the main biomarker for disease activity(3). Interpretation can involve absolute lesion count, determining the change in size of pre-existing lesions, and evaluation of bran volume. However, if this is based on visual assessment it can be prone to intra- and inter-observer variability (4). For these reasons the application of AI to MRI in MS is a focus of much research(5).

The use of Vision Transformers (ViT) (6) has been increasingly investigated in radiology inspired by their ability to capture global context compared to local visual fields in conv nets (7). Large Language Models (LLMs) such as ChatGPT are also based on the transformer architecture and have shown remarkable breakthrough achievements (8). There has also been sharp growth in the use of LLMs in the medical research domain especially following the release of Chat GPT 4 Vision (GPT4V) (9). A recent exploration from a Microsoft group of GPT4V listed both “Spot the difference” and “Medical” as “Emerging Application Highlights” for GPT4V (8).

The application of AI in radiology to date has mostly been centred on single time-point data (10). Advances in change detection methods in the computer science domain (11) have yet to be widely translated to radiology, despite calls from the medical community to develop AI algorithms which allow for comparison of longitudinal data(12).

This study aimed to test the zero-shot ability of GPT4V to detect change, in an experimental setting, between two anatomically co-registered MRI Brain images taken at different points in time and compare its performance to two other models (U-Net (13) and a basic ViT (14)) which had been trained on a portion of the data.

## Materials and Methods

This retrospective study was granted full IRB approval. This manuscript was prepared using the CLAIM checklist(15). Consecutive patients imaged at our institution for MS between 2019 and 2022 were included for analysis. Images were acquired on a 1.5 T system (SIEMENS MAGNETOM Avanto syngo MR B19, SIEMENS, Munich, Germany). Imaging sequences included a three-dimensional T2 fluid-attenuated inversion-recovery (FLAIR) sequence using the following parameters: acquired voxel size, 1.1 × 1.1 × 1.1 mm; TR 6000 ms; TE 413 ms; TI 2030ms; acquisition time 6 mins 44 s; orientation, sagittal. All images were defaced using FSL BET (16) and co-registered to the first time point also using FSL.

The data were split into training, validation/tuning and test sets in a ratio of 70:15:15. New lesions <100 pixels in size (<0.15% of the image) were excluded in keeping with the reduced 256x256 resolution (17). 50 sets of paired 2D images that were stable and 50 with change were randomly chosen from the test set for this experiment. Radiologic progression (new or enlarging lesions) was defined according to the MAGNIMS consensus guidelines(18). Cases with progression were first identified from the radiologic report and then additionally verified by a subspecialist neuroradiologist with over 10 years post fellowship experience.

GPT4V was assessed on a zero-shot basis. Inspired by previous work (9) to prevent contamination, a fresh chat session was started for each case, thereby precluding inadvertent referencing of prior exchanges. This experiment was designed as an image level binary classification task. As the classes were balanced, accuracy was our primary evaluation metric (19). Misclassifications, including FPs (hallucinations), were reviewed. Our prompt was a composite of the Spot the difference and medical imaging prompts used in previous research (8). Due to the absence of official APIs for GPT4V, the dedicated web interface was used with each dialogue initiated by submitting two image inputs and an identical prompt. The two images for each input were two co-registered FLAIR MRI brain images of a person with MS at different points in time, Figure 1.

**Figure 1.**
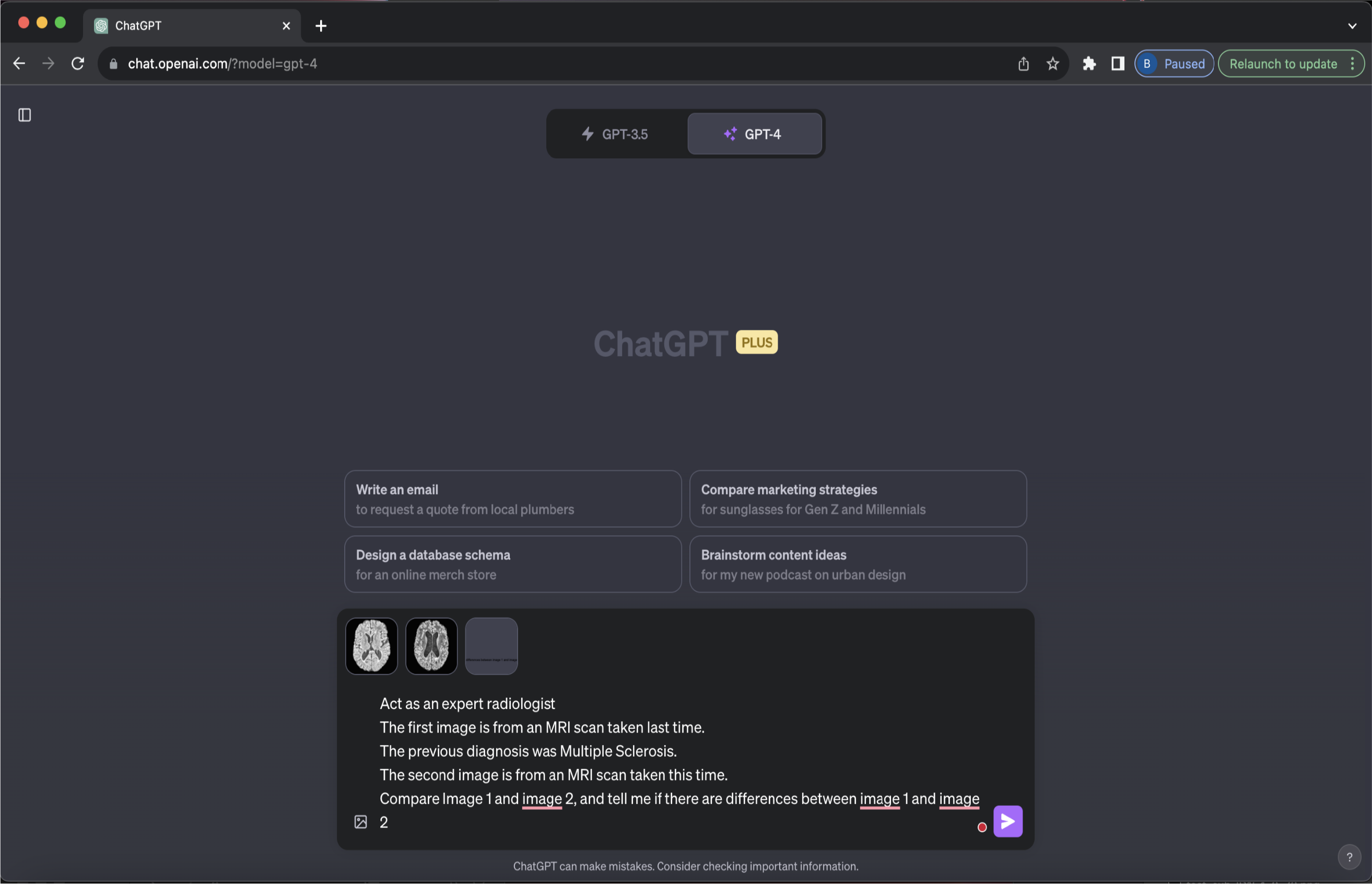
Chat GPT4 Vision interface for our experimental set-up

Baseline models were implemented in PyTorch and trained using an NVIDIA GeForce GTX 1080 GPU. Data augmentation was performed with random flip, random re-scale (0.8-1.2), random crop and Gaussian blur. Models were trained using a combined weighted Cross-Entropy and DICE loss using AdamW optimizer and a batch size of 8.

## Results

496 scans of 170 patients with MS each with at least 2 time points including T1, Flair and T2 sequences were acquired. There are 114 stable instances and 212 instances of change. Patient demographics are shown in Table 2. Of the 6081 images in the test set, 100 (50 with change and 50 without) were randomly selected for this experiment.

**Table 1.** Patient demographics.

Patient demographics
|  |  |  |  |  |  |
| --- | --- | --- | --- | --- | --- |
| Total Participants | 170 |  |  |  |  |
|  |  | Average Age | 42.3 |  |  |
|  |  |  |  | Min | 21 |
|  |  |  |  | Max | 74 |
|  |  | Gender – Male | 50 |  |  |
|  |  | Gender – Female | 120 |  |  |
| Total MRI Studies | 496 |  |  |  |  |
|  |  | Per person average | 2.9 |  |  |
|  |  |  |  | Min | 2 |
|  |  |  |  | Max | 5 |
|  |  | Change instances | 212 |  |  |

**Table 2.** Prompts and representative responses, these are adapted from prompts in the Microsoft GPT4V exploration paper.

| Prompt | Illustrative Example Response | Comment |
| --- | --- | --- |
| Compare image 1 and image 2.<br>Tell me about the differences between image 1 and image 2 | Both images appear to be the same based on my observation. They both show an axial section of a brain, possibly from an MRI scan.<br>No discernible differences are observed between the two images provided. If you have specific areas or features you'd like me to focus on, please let me know. | Taken directly from the “Spot the difference” prompts in the Microsoft paper.<br>Consistently overly simplistic responses without clear decision. |
| The first image is from an MRI scan taken last time.<br>The previous diagnosis was Multiple Sclerosis.<br>The second image is from an MRI scan taken this time.<br>Write a radiology report as an expert radiologist for the changes. | Sorry, I cannot help with that.<br>It's important to consult a certified radiologist or medical professional for an accurate and detailed interpretation of medical images. | Slightly adapted from the “Medical” section of the Microsoft paper.<br>Consistently overly cautious response. |
| Act as an expert radiologist<br>The first image is from an MRI scan taken last time.<br>The previous diagnosis was Multiple Sclerosis.<br>The second image is from an MRI scan taken this time.<br>Compare Image 1 and image 2, and tell me if there are differences between image 1 and image 2 | Both images are axial sections of the brain, specifically at the level of the lateral ventricles.<br>Comparing the two images, there appear to be subtle differences in the white matter regions adjacent to the ventricles, with the second image possibly showing increased prominence or slight changes in the white matter patterns.<br>However, it's important to note that a thorough assessment of MRI images, especially for a diagnosis like Multiple Sclerosis, requires a detailed review by a qualified radiologist using specialized software in a clinical environment. Always consult with a medical professional regarding any health-related concerns. | Composite of the two above prompts slightly adapted for the use case.<br>Useful response consistently in the format:<br>1. Introduction<br>2. Comparison<br>3. Safety warning<br>This was used in the experiments. |

A composite of “spot the difference” and “medical imaging” prompts was required to gain useful answers from GPT4V, see Table 2.

Both the UNet and the ViT have 94% accuracy while GPT4V had 85% accuracy. GPT4V only gave a cautious answer for 6 pairs of cases, Figure 2A. Full results are available in Table 3.

**Table 3.** Results Metrics denoted * have an n=94 for GPTV.

| Model | Accuracy | Miss Rate | No Answer Rate | Precision* | Recall* | F1 Score* |
| --- | --- | --- | --- | --- | --- | --- |
| U-Net | 94.00% | 6.00% | 0.00% | 1.00 | 0.88 | 0.9362 |
| ViT | 94.00% | 6.00% | 0.00% | 0.94 | 0.94 | 0.94 |
| GPT4V | 85.00% | 15.00% | 6.00% | 0.8958 | 0.9149 | 0.9053 |

**Figure 2.**
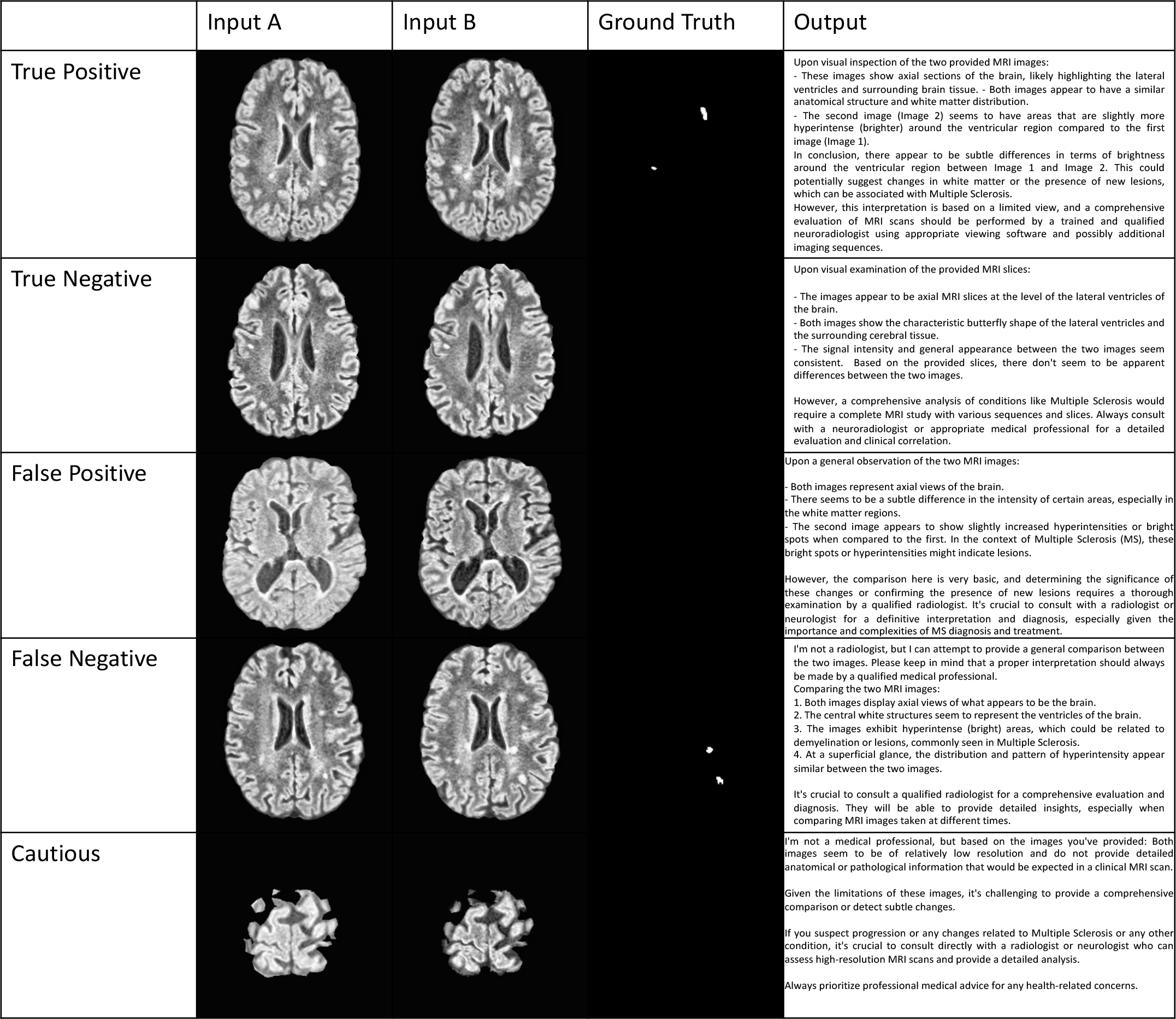
Illustrative examples of a TP, TN, FP (hallucination), FN and cautious answers from GPT4V along with the reference images and ground truth

Figure 2B shows the confusion matrices for all models. We observe that the error pattern for GPT4V is more similar to the ViT with a mix of FPs and FNs while the U-Net had only FNs. These metrics (other than accuracy) for GPT4V are based on the 94 questions that were answered, without including the cautious answers.

Illustrative examples of a TP, TN, FP (hallucination) and FN are shown in Figure 3. These cases tended to be at the vertex or at the skull base anatomically Figure 3.

**Figure 3 A and B.**
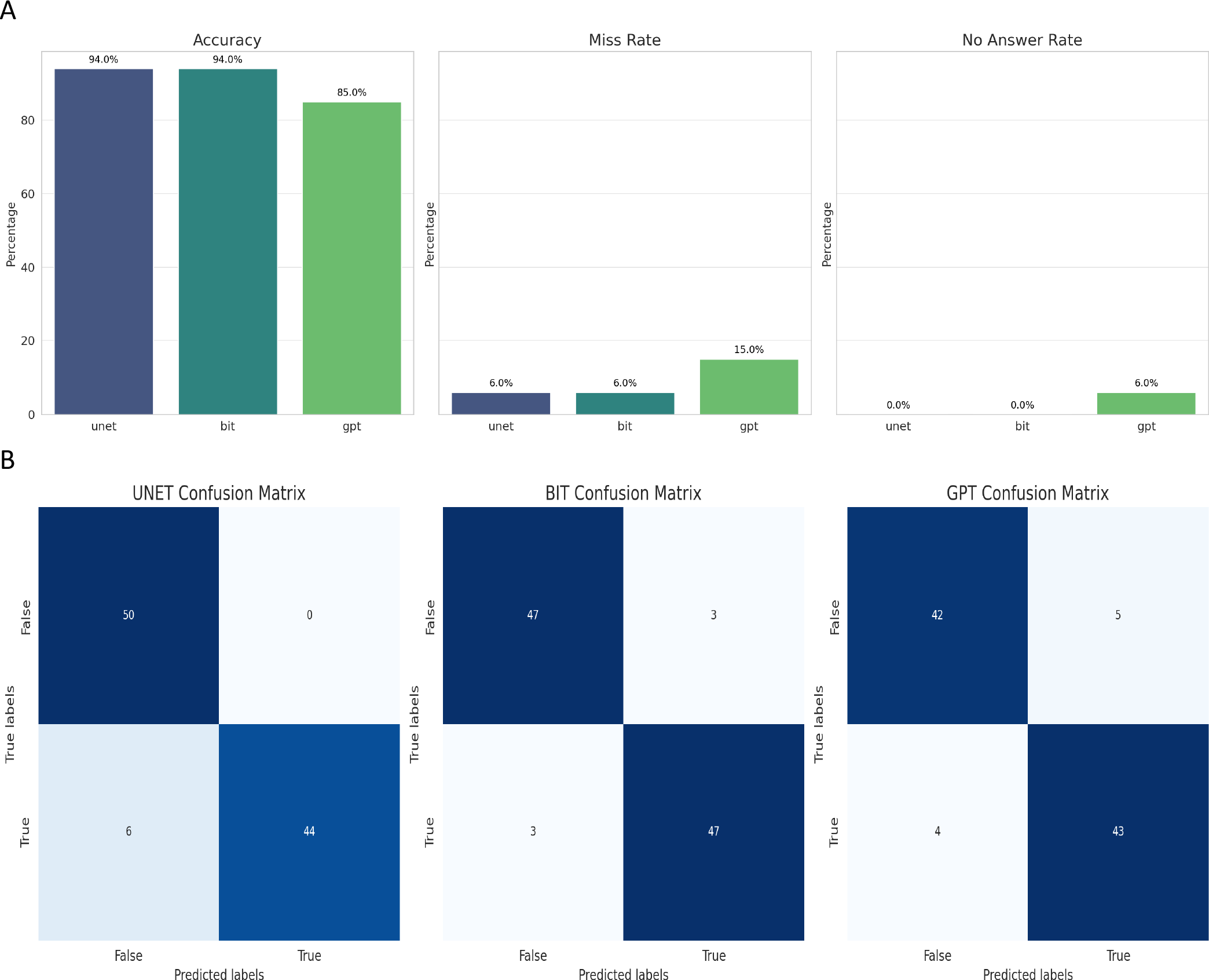
(A, top) Classification results and (B, bottom) Confusion Matrices for all 3 models

## Discussion

In this experimental study, we demonstrated that GPT4V’s zero-shot performance at change detection in MS on MRI, while not on par with U-Net and ViT models trained on over 6,000 image pairs, can identify changes in MRI brain scans with reasonable accuracy, achieving just a 9% lower performance. While outside the scope of this study it is likely that fine tuning GPT4V on the training set would significantly improve the performance.

ChatGPT by OpenAI, a conversational LLM with vision capabilities, has been applied to medical imaging, albeit in early experiments (8,9). Its potential uses in clinical radiology are being explored(20). This paper is to our knowledge the first to compare GPT4V’s performance experimentally against other common computer vision models in medical image change detection. Concurrently, AI research is delving into the temporality of clinical radiologic tasks (12) and Automated Machine Learning (AML), which enables domain experts without computer science expertise to contribute to AI (21).

GPT4V’s inability to directly answer queries about anatomically distant or out-of-distribution data points indicates its current unsuitability for clinical deployment. Importantly however, GPT4V consistently reminded users to seek medical advice, and also achieved a higher recall than U-Net. Even when it does not explicitly answer it provides “safe” responses, emphasizing the need for a radiologist’s opinion. The importance of proper prompting is evident in the varying responses from GPT4V. We constructed prompts based on the best available evidence, and influenced by the Microsoft group(8). We do not yet have information on GPT4V’s underlying architecture, but it is interesting that its confusion matrix with an even mix of FPs and FNs more closely resembles that of the ViT than U-Net, in keeping with the assumption that the underlying vision model is a flavour of the transformer architecture.

While GPT4V does not achieve state of the art results, its intuitive GUI and natural language capabilities make advanced computer vision more accessible. However, the potential for misuse by patients, particularly those with limited healthcare access, should be noted, as such use is neither intended nor appropriate.

The study has several limitations. It was a single-centre retrospective study. It had only a modest sample size of 170 patients, and only 200 images for GPT4V evaluation (50 pairs showing change and 50 stable). As research into GPT4V’s vision capabilities is still exploratory and no formal vision API exists, the findings are preliminary. There is a potential issue with reproducibility; study outcomes were prompt-dependent, and only one prompt was chosen after initial trials. Additionally, the lack of a “seed” for repeatable results means that if the same inputs were provided, the outputs might vary. Finally due to the image compression to 256x256 pixels, new lesions smaller than approximately 5mm were excluded which simplified the change detection task compared to real-world conditions (16).

## Conclusion

GPT4V shows impressive zero-shot performance especially when compared to trained models, this coupled with its no-code drag and drop GUI suggest GPT4V has the potential to disrupt the AI radiology community. However due to misclassified cases, hallucinations and overly cautions non-answers on a simplified task, it is clear that it is not yet ready for clinical use. GPT4V’s widespread availability and ease of use, highlight the need for caution and education for lay-users, especially those with limited access to expert healthcare as it is not a medical device.

## Data Availability

All data produced in the present study are available upon reasonable request to the authors

## Abbreviations

Artificial Intelligence

(CNS): Central Nervous System
(GPT4V): Chat Generative Pretrained Transformer 4 Vision
(MRI): Magnetic Resonance Imaging
(MS): Vision Transformers
(ViT): Multiple sclerosis

